# Surface EEG to identify cognitive motor dissociation after acute brain injury

**DOI:** 10.64898/2025.12.16.25342313

**Authors:** Satoshi Egawa, Nicole Casson, Joel Neves Briard, Qi Shen, Vedant Kansara, Itamar Niesvizky-Kogan, Elizabeth Carroll, Jerina C. Carmona, You Lim Song, Alex J. Klein, Angela Velazquez, Wells Andres, Shivani Ghoshal, David Roh, Sachin Agarwal, Soojin Park, E Sander Connolly, Jan Claassen

**Affiliations:** Department of Neurology, Columbia University Medical Center, New York, NY, USA; NewYork-Presbyterian Hospital, New York, NY, USA; Department of Biomedical Informatics, Columbia University Medical Center, New York, NY, USA; Department of Neurosurgery, Columbia University Medical Center, New York, NY, USA

**Keywords:** EEG, ACNS criteria, CMD, covert consciousness, cognitive motor dissociation, coma

## Abstract

**Objective:** Cognitive motor dissociation (CMD) is associated with long-term recovery in acute brain injury, but CMD testing is only available in few centers. Our objective was to identify surface EEG patterns with high sensitivity or positive predictive value (PPV) for CMD in patients with acute disorders of consciousness to refine allocation of this resource-intensive test.

**Methods:** In this observational cohort study, we enrolled clinically unresponsive, acutely brain injured patients who underwent continuous surface EEG and CMD assessments. CMD was detected by applying a machine learning algorithm to EEG acquired during a motor command paradigm presentation.

Electroencephalographers blinded to CMD test results applied standardized ACNS criteria to the EEGs acquired during CMD assessments. We calculated accuracy measures of surface EEG findings for CMD test results using generalized estimating equations, with an exchangeable matrix and accounting for repeated measures per patient.

**Results:** We included 185 patients (mean age: 62 ± 17; 85 [46%] female) and 282 CMD assessments. CMD testing was positive in 39 (14%) assessments. Sensitivity and PPV of normal background voltage, symmetry and continuity were respectively 77% (95%-CI: 60-88%) and 19% (95%-CI: 13-26%), 74% (95%-CI: 58-86%) and 14% (95%-CI: 10-20%), and 74% (95%-CI: 58-86%) and 14% (95%-CI: 9-19%). All EEGs with burst suppression, suppression, sporadic epileptiform discharges, lateralized periodic discharges, bilateral independent periodic discharges, electrographic seizures and brief potentially ictal rhythmic discharges had negative CMD tests.

**Interpretation:** Surface EEG findings are not reliable to screen for CMD or to identify patterns conferring higher CMD pretest probability.

## INTRODUCTION

Impairment of consciousness is common after acute brain injury and carries a tremendous emotional and socioeconomic burden for families and health care systems.^1^ Accurate and early prediction of recovery after brain injury is challenging, as delayed recovery can occur when prolonged supportive care is provided. In acutely brain injured patients, subtle signs of consciousness, such as those detected in minimally conscious states, have been shown to herald later further neurological recovery.^2, 3^ Moreover, brain activation to motor commands has been detected in 14-17% of critically-ill, clinically unresponsive patients using functional MRI and EEG-based machine learning algorithms.^4, 5^ This finding, termed cognitive-motor dissociation (CMD), suggests these patients are able to participate in mental activities despite appearing to be unresponsive on behavioral assessments, indicating some level of preserved consciousness.^5, 6^ Recently, CMD has been robustly associated with long-term recovery of independence in this patient population.^7, 8^

Despite the potential implications of CMD in prognostication of acute disorders of consciousness (DoC), its widespread application has been limited by a lack of technical expertise, and the complexity of data acquisition and analysis.^9, 10^ There are currently no reliable methods to estimate CMD pretest probability in order to optimize the allocation of this resource-intensive test. We hypothesized that certain surface EEG patterns have very high sensitivity for CMD whereas others have very high positive predictive value for CMD, such that these patterns may respectively aid in screening for CMD and in allocating CMD testing to individuals at high CMD pretest probability. The objective of this study was therefore to identify surface EEG patterns with high sensitivity or positive predictive value for CMD in patients with acute DoC to refine allocation of CMD testing.

## METHODS

### Patients and design

This is a cross-sectional analysis nested in a prospective cohort study. The study population comprises adult patients who are unresponsive to verbal commands following an acute and severe brain injury. From July 2014 through January 2022, we prospectively screened all patients admitted with acute brain injury to the Columbia University Irving Medical Center neuroscience intensive care unit (ICU) within 3 days after admission for inclusion in our cohort study. Patients were screened for the absence of the ability to follow simple spoken commands — for example, “stick out your tongue” or “show me two fingers with your right hand.”^7^ We enrolled consecutive patients who (1) were determined by bedside Coma Recovery Scale-revised (CRS-R) assessment to be in a coma, vegetative state/unresponsive wakefulness syndrome (VS/UWS), or minimally conscious state (MCS)-minus (defined as unresponsiveness with preserved visual fixation, visual pursuit, or localization to noxious stimuli); (2) were unable to follow spoken commands (including assessments for mimicking);^11^ (3) were undergoing or were expected to undergo continuous EEG monitoring as part of their clinical care; and (4) spoke English or Spanish as their native language. Patients were excluded that were (1) less than 18 years old, (2) pregnant, (3) had a pre-existing DoC before the onset of acute brain injury prompting hospital admission, (4) had confounding neurological conditions (i.e., advanced dementia), (5) deafness prior to acute brain injury, (6) uncontrollable seizures (i.e., refractory status epilepticus), (7) infection with COVID-19, (9) clinical recovery of the ability to follow commands prior to enrollment, (10) logistical reasons hindering enrollment, (11) very high level of sedation (i.e., pentobarbital anesthesia for refractory elevated intracranial pressure), and (12) surrogate not giving consent.

### Study oversight

This study was approved by the Columbia University Irving Medical Center local institutional review board. Written informed consent was obtained from patients’ surrogates. All patients who recovered consciousness and demonstrated decision-making capacity were given the opportunity to withdraw from the study. Study reporting follows STARD guidelines.^12^

### Behavioral assessments

As part of our cohort study research protocol, we performed serial assessments of consciousness using the CRS-R throughout the ICU stay, including prior to each CMD assessment. Each assessment result was categorized as coma, VS/UWS, MCS-, MCS+, and emergence from MCS (EMCS).

### EEG recording

As per clinical protocol in our ICU, all patients with impairment of consciousness and acute brain injury underwent at least 2 days of continuous EEG monitoring for the primary purpose to detect electrographic seizures. EEG monitoring was obtained applying a standard 21-electrode montage placed according to the International 10-20 system^13^ using a digital bedside video EEG monitoring system (XLTEK, Excel-Tech Corp., Natus Medical Incorporated, Oakville, Ontario, Canada; low-pass filter = 70 Hz, high-pass filter = 1 Hz, sampling rate = 200 Hz).

### Sedation

Sedation was documented at the time of EEG recordings and categorized into five groups: none for those not receiving any sedation; minimal, for those receiving pushes of sedative medications only; low, for those receiving continuously administered doses of midazolam at 0.15 mg/kg/h or less, propofol at 4 mg/kg/h or less, fentanyl at 2 µg/kg/h or less, or any dose of dexmedetomidine; moderate, for those receiving continuously administered doses of midazolam above 0.15 mg/kg/h, propofol above 4 mg/kg/h, fentanyl above 2 µg/kg/h, or any dose of ketamine; and high, for those receiving barbiturate infusions. We did not include any patients that received high level sedation. Whenever possible and safe, clinicians weaned sedation prior to behavioral and CMD assessments.

### Reference standard: CMD testing

The reference standard in this analysis was the CMD test. As previously published, the test comprises an EEG-based motor command paradigm and a machine learning algorithm.^7^ In summary, during each CMD assessment, patients were exposed to 6 blocks of spoken command instructions alternating between “keep opening and closing your right hand” and “stop opening and closing your right hand.” We recorded three blocks in which the patient was asked to move the right hand and three blocks in which the patient was asked to move the left hand. The total duration of the motor command paradigm was approximately 25 minutes. The EEG acquired during the paradigm was analyzed by calculating spectral power in predefined frequency ranges for each electrode. A support vector machine with a linear kernel was applied to the spectral power during the “opening” and “closing” commands to detect brain activation to motor commands.^7, 8^ CMD test results were dichotomized as positive or negative.

### Index test: surface EEG

The index test in this analysis was the qualitative interpretation of the surface EEG acquired during the motor command paradigm. We expected patients included in this study would have many days of EEG monitoring and a variable number of CMD assessments: for patients that had >2 CMD assessments available, we therefore selected two EEGs using a random number generator. An experienced electroencephalographer and neurointensivist (SE) blinded to the clinical course, CMD test results, and clinical outcomes annotated surface EEGs using the 2021 ACNS Standardized Critical Care EEG Terminology.^14^ The following findings were scored: background activity (voltage, dominant frequency, symmetry, continuity, posterior dominant rhythm [PDR], reactivity, state changes, cyclic alternating pattern of encephalopathy [CAPE], and presence of an anterior-posterior [AP] gradient), epileptiform activity (sporadic epileptiform discharges [SED], rhythmic periodic patterns [RPPs], generalized periodic discharges [GPDs], lateralized periodic discharges [LPDs], bilateral independent periodic discharges [BIPDs], unilateral independent periodic discharges [UIPDs], multifocal periodic discharges [MfPDs], generalized rhythmic delta activity [GRDA], lateralized rhythmic delta activity [LRDA], generalized spike and wave [GSW], lateralized spike and wave [LSW], electrographic seizures and status epilepticus, brief potentially ictal rhythmic discharges [BIRDs] and ictal-interictal continuum [IIC]). EEG findings were dichotomized as present or absent.

### Statistical analyses

Categorical variables were reported as counts (percentages), and continuous variables as means (standard deviation) or medians (interquartile range), as appropriate. We built chord diagrams to illustrate the co-occurrence of background activity characteristics (voltage, symmetry and continuity) in EEGs, stratifying by CMD test result.^15^ In primary analyses, we calculated the sensitivity, specificity, positive predictive values (PPV) and negative predictive values (NPV) of EEG findings for CMD test results using generalized estimating equations, with an exchangeable correlation matrix and accounting for repeated measures per patient. In the context of this analysis, sensitivity represents the proportion of positive CMD tests that have the EEG finding of interest, and specificity represents the proportion of negative CMD tests that do not have the EEG finding of interest. The PPV represents the probability that a surface EEG with the finding of interest has a positive CMD test. Therefore, a surface EEG finding’s PPV provides an estimate of CMD pretest probability when the finding is present. The NPV represents the probability that a surface EEG without the finding of interest has a negative CMD test. In secondary analyses, we evaluated the interrater reliability of surface EEG scoring. Two experienced electroencephalographers (SE, EC) that were blinded to each other’s interpretations independently rated a random sample of 20 included surface EEGs according to ACNS criteria. We calculated concordance ratios (Cohen’s kappa) per finding category (symmetry, frequency, continuity, etc.); values less than 0 signify poor concordance, 0 to 0.20 slight, 0.21 to 0.40 fair, 0.41 to 0.60 moderate, 0.61 to 0.80 substantial, and 0.81 to 1.0 almost perfect agreement.^16^ No hypothesis testing was required in this analysis. Analyses were performed using R (version 4.0.3).

## RESULTS

Among 655 patients screened for inclusion, 185 met the study selection criteria (**Supplementary Figure 1** and **Supplementary Table 1**). Common reasons for exclusion were that patients started following commands prior to enrollment (n=148), logistical reasons preventing assessments (n=120), no continuous EEG (n=69), confounding neurological conditions (n=54), and uncontrollable seizures (n=35). Included patients had a mean age of 62 ± 17 years; 85 (46%) were female (**Table 1**). Brain injury etiologies were intracerebral hemorrhages in 65 (35%), cardiac arrest in 34 (18%), subarachnoid hemorrhage in 27 (15%), traumatic brain injury or subdural hematoma in 24 (13%), acute ischemic stroke in 13 (7%) and other etiologies in 22 (12%) patients. At admission, the median admission Glasgow Coma Scale score was 6 (3-8) and the median CRS-R score was 3 (1-5). Among the 185 included patients, we collected 282 behavioral-EEG-CMD assessments. Behavioral assessments were consistent with coma in 142 (50%), VS/UWS in 70 (25%), and MCS-minus in 70 (25%).

**Table 1.** Patient cohort characteristics (N=185) ^a^.

|  |  |  |
| --- | --- | --- |
| Demographics |  |  |
| Age — years |  | 62 (17) |
| Sex, female |  | 85 (46) |
| Admission clinical characteristics |  |  |
| Glasgow Coma Scale score |  | 6 [3-8] |
| Etiology of acute brain injury |  |  |
| Intracerebral hemorrhage |  | 65 (35) |
| Cardiac arrest |  | 34 (18) |
| Subarachnoid hemorrhage |  | 27 (15) |
| Traumatic brain injury or subdural hematoma |  | 24 (13) |
| Acute ischemic stroke |  | 13 (7) |
| Other <sup>b</sup> |  | 22 (12) |
| Behavioral assessments, in-hospital |  |  |
| CRS-R, median at admission [IQR] |  | 3 [1-5] |
| CRS-R, median of the worst [IQR] |  | 1 [0-3] |
| CRS-R, median of the best [IQR] |  | 3 [1-9] |
| Cognitive motor dissociation (CMD) positive <sup>c</sup> |  | 28 (15) |

<sup>a</sup> Data reported as n (%), mean (standard deviation), or median [interquartile range] as appropriate.
<sup>b</sup> Other etiologies include brain tumor, encephalitis, encephalopathy and status epilepticus.
<sup>c</sup> A patient was considered CMD positive if at least one of their CMD tests was positive.
CRS-R denotes Coma Recovery Scale-Revised.

### Surface EEG findings

Among the 185 included patients, 88 contributed a single surface EEG and 97 patients contributed two EEGs (baseline characteristics for patients contributing one versus two recordings are provided in **Supplementary Table 2**). Among 282 surface EEGs, 163 (58%) recordings were made while patients were not receiving any sedation, 13 (5%) with minimal, 81 (29%) with low, 19 (7%) with moderate and 0 with heavy sedation (sedation data was undetermined in 6 patients). Surface EEG findings in the 282 records are detailed in **Table 2**. Background voltage was normal in 162 (57%), with a dominant background frequency in the theta range in 137 (49%), alpha in 70 (25%), delta in 47 (17%), and beta in 2 (1%) recordings. EEG background was symmetric in 198 (70%), whereas it had mild and marked asymmetry in 39 (14%) and 45 (16%), respectively. Background activity was continuous in 212 (75%) EEGs, nearly continuous in 21 (7%), discontinuous in 23 (8%), burst suppression in 10 (4%) and suppression in 16 (6%). A PDR was detected in 93 (33%) EEGs, 177 (63%) were categorized as reactive, state changes were present in 174 (62%), and an AP gradient was seen in 10 (4%). Sporadic epileptiform discharges were observed in 5 (2%) of EEGs. Overall, 85 (30%) EEGs had rhythmic and periodic patterns: GPDs, LPDs, BIPDs, GRDA, and LRDA were observed in 41 (15%), 17 (6%), 2 (1%), 21 (7%), and 4 (1%) recordings, respectively. Three (1%) EEGs had electrographic seizures, 5 (2%) had BIRDs and 35 (12%) had findings on the IIC. No EEGs had UIPDs, MfPD, GSW, LSW, electroclinical seizures, electrographic status epilepticus or electroclinical status epilepticus.

**Table 2.** Surface EEG findings across 282 assessments ^a^.

|  | Entire EEG sample<br>(n=282) | CMD-positive EEGs (n=39) | CMD-negative EEGs (n=243) |
| --- | --- | --- | --- |
| **EEG background** |  |  |  |
| Normal background voltage ($\geq 20 \mu V$) | 162 (57) | 30 (77) | 132 (54) |
| Dominant background frequency |  |  |  |
| Beta | 2 (1) | 0 (0) | 2 (1) |
| Alpha | 70 (25) | 13 (33) | 57 (24) |
| Theta | 137 (49) | 20 (51) | 117 (48) |
| Delta | 47 (17) | 5 (13) | 42 (17) |
| Symmetry |  |  |  |
| Symmetric | 198 (70) | 29 (74) | 169 (70) |
| Mild asymmetry | 39 (14) | 2 (5) | 37 (15) |
| Marked asymmetry | 45 (16) | 8 (21) | 37 (15) |
| Continuity |  |  |  |
| Continuous | 212 (75) | 29 (74) | 183 (74) |
| Nearly continuous | 21 (7) | 3 (8) | 18 (8) |
| Discontinuous | 23 (8) | 7 (18) | 16 (7) |
| Burst suppression | 10 (4) | 0 (0) | 10 (4) |
| Suppression | 16 (6) | 0 (0) | 16 (7) |
| Posterior dominant rhythm | 93 (33) | 15 (39) | 78 (32) |
| Reactivity | 177 (63) | 25 (64) | 152 (63) |
| State change | 174 (62) | 27 (69) | 147 (61) |
| AP gradient | 10 (4) | 1 (3) | 9 (4) |
| Cyclic alternating pattern of encephalopathy | 25 (9) | 7 (18) | 18 (7) |
| **Epileptiform activity <sup>b</sup>** |  |  |  |
| Sporadic epileptiform discharges | 5 (2) | 0 (0) | 5 (2) |
| Rhythmic and periodic patterns | 85 (30) | 11 (28) | 74 (30) |
| Generalized periodic discharges | 41 (15) | 7 (18) | 34 (14) |
| Lateralized periodic discharges | 17 (6) | 0 (0) | 17 (7) |
| Bilateral independent periodic discharges | 2 (1) | 0 (0) | 2 (1) |
| Generalized rhythmic delta activity | 21 (7) | 3 (8) | 18 (7) |
| Lateralized rhythmic delta activity | 4 (1) | 1 (3) | 3 (1) |
| **Seizures, BIRDS and IIC <sup>b</sup>** |  |  |  |
| Electrographic seizures | 3 (1) | 0 (0) | 3 (1) |
| Brief potentially ictal rhythmic discharges (BIRDS) | 5 (2) | 0 (0) | 5 (2) |
| Ictal-interictal continuum (IIC) | 35 (12) | 1 (3) | 34 (14) |
| Periodic discharges or spike-wave patterns >1.0 Hz but <2.5 Hz over 10 seconds | 28 (10) | 1 (3) | 27 (11) |
| Periodic discharges or spike-wave patterns >0.5 Hz and <1 Hz over 10 seconds with a plus modifier or fluctuation | 6 (2) | 0 (0) | 6 (3) |
| Lateralized rhythmic delta activity >1 Hz for at least 10 seconds with a plus modifier or fluctuation | 1 (0) | 0 (0) | 1 (0) |

<sup>a</sup> Data reported as n (%).<sup>b</sup> No EEGs had unilateral independent periodic discharges, multifocal periodic discharges, generalized spike and wave, lateralized spike and wave, electroclinical seizures, electrographic status epilepticus or electroclinical status epilepticus.

### Accuracy of surface EEG for CMD

CMD tests were positive in 39 (14%) and negative in 243 (86%) assessments. Sensitivity and specificity of surface EEG findings for CMD are detailed in **Figures 1-2**, and PPV and NPV are provided in **Figures 3-4**. Normal background voltage had a sensitivity of 77% (95% CI: 60-88%), a specificity of 46% (95% CI: 39-53%) and a PPV of 19% (95% CI: 13-26%). Symmetric background had a sensitivity of 74% (95% CI: 58-86%), a specificity of 31% (95% CI: 25-38%) and a PPV of 14% (95% CI: 10-20%). Continuous background had a sensitivity of 74% (95% CI: 58-86%), a specificity of 25% (95% CI: 19-32%) and a PPV of 14% (95% CI: 9-19%). Presence of a PDR had a sensitivity of 48% (95% CI: 31-66%), a specificity of 62% (95% CI: 54-69%), and a PPV of 15% (95% CI: 9-25%). Reactivity has a sensitivity of 86% (95% CI: 68-95%), a specificity of 19% (95% CI: 13-26%), and a PPV of 14% (95% CI: 10-20%). State change had a sensitivity of 69% (95% CI: 53-81%), a specificity of 39% (95% CI: 33-47%), and a PPV of 15% (95% CI: 10-22%). AP gradient had a sensitivity of 3% (95% CI: 0-16%), a specificity of 96% (95% CI: 92-98%) and a PPV of 10% (95% CI: 1-48%). Several abnormal background features and epileptiform activity findings had low sensitivity (**Figure 1**) and high specificity for CMD (**Figure 2**). No EEG finding had a high PPV for CMD, and most PPV estimates had very wide confidence intervals (**Figure 3**). All EEGs with burst suppression or suppression, SEDs, LPDs, BIPDs, electrographic seizures and BIRDs had negative CMD tests. EEG findings had overall similar NPV (**Figure 4**).

**Figure 1.**
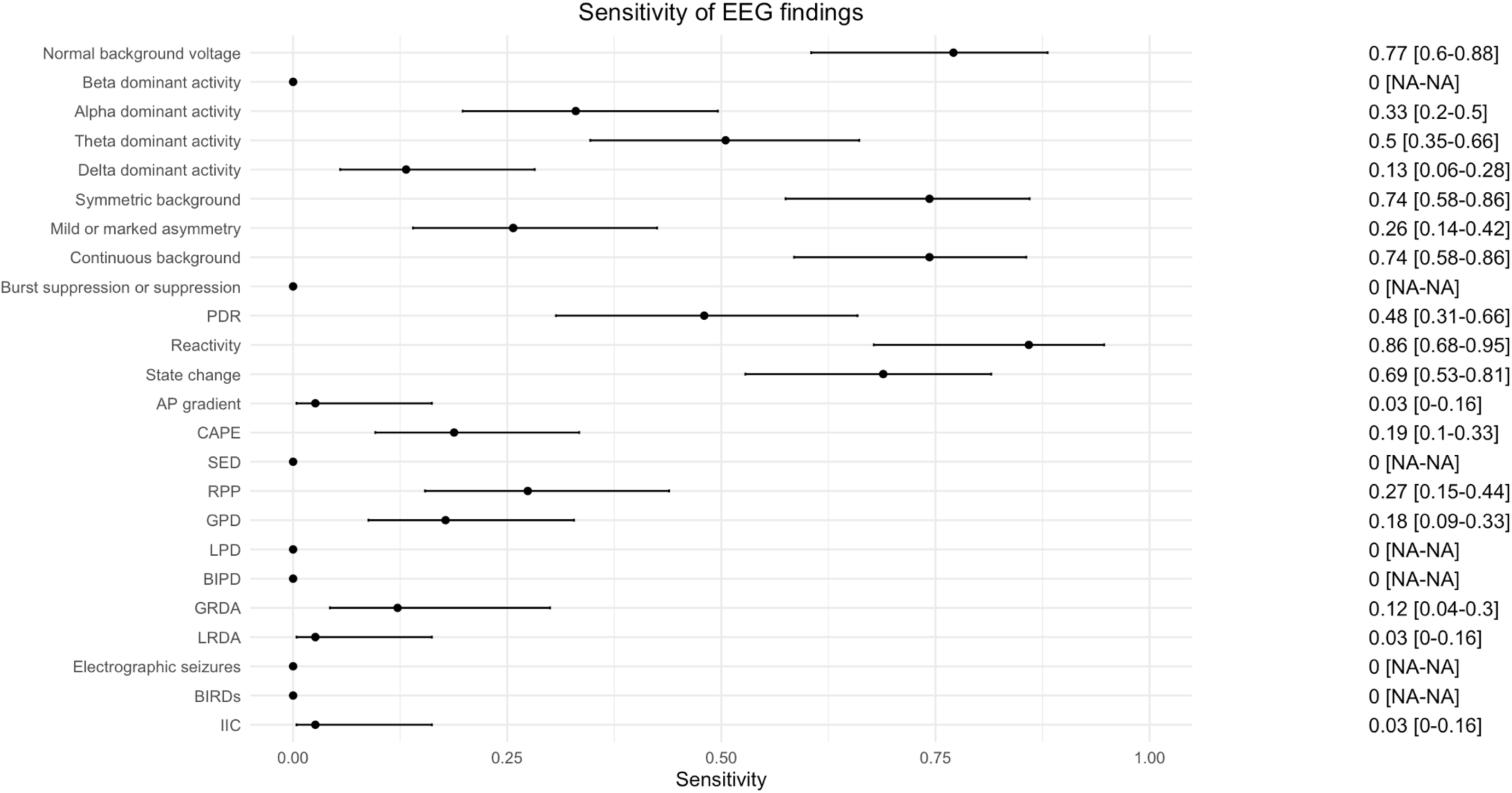
Sensitivity of surface EEG findings for cognitive-motor dissociation. Bars represent 95% confidence intervals. Confidence intervals were not calculable (NA) for the sensitivity of beta dominant activity, burst suppression or suppression, SED, LPD, BIPD, electrographic seizures and BIRDs due to no observed true positives.

**Figure 2.**
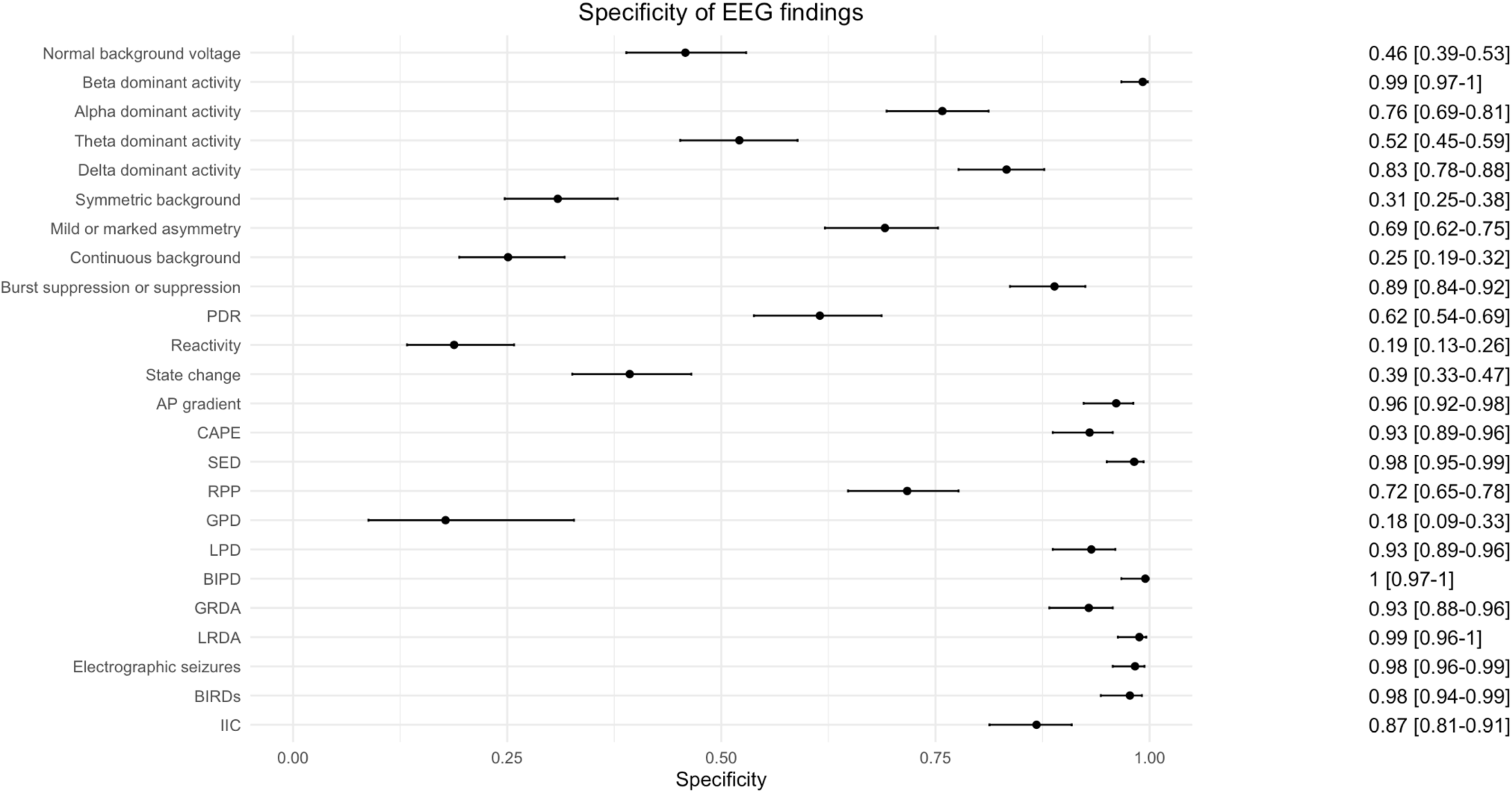
Specificity of surface EEG findings for cognitive-motor dissociation. Bars represent 95% confidence intervals.

**Figure 3.**
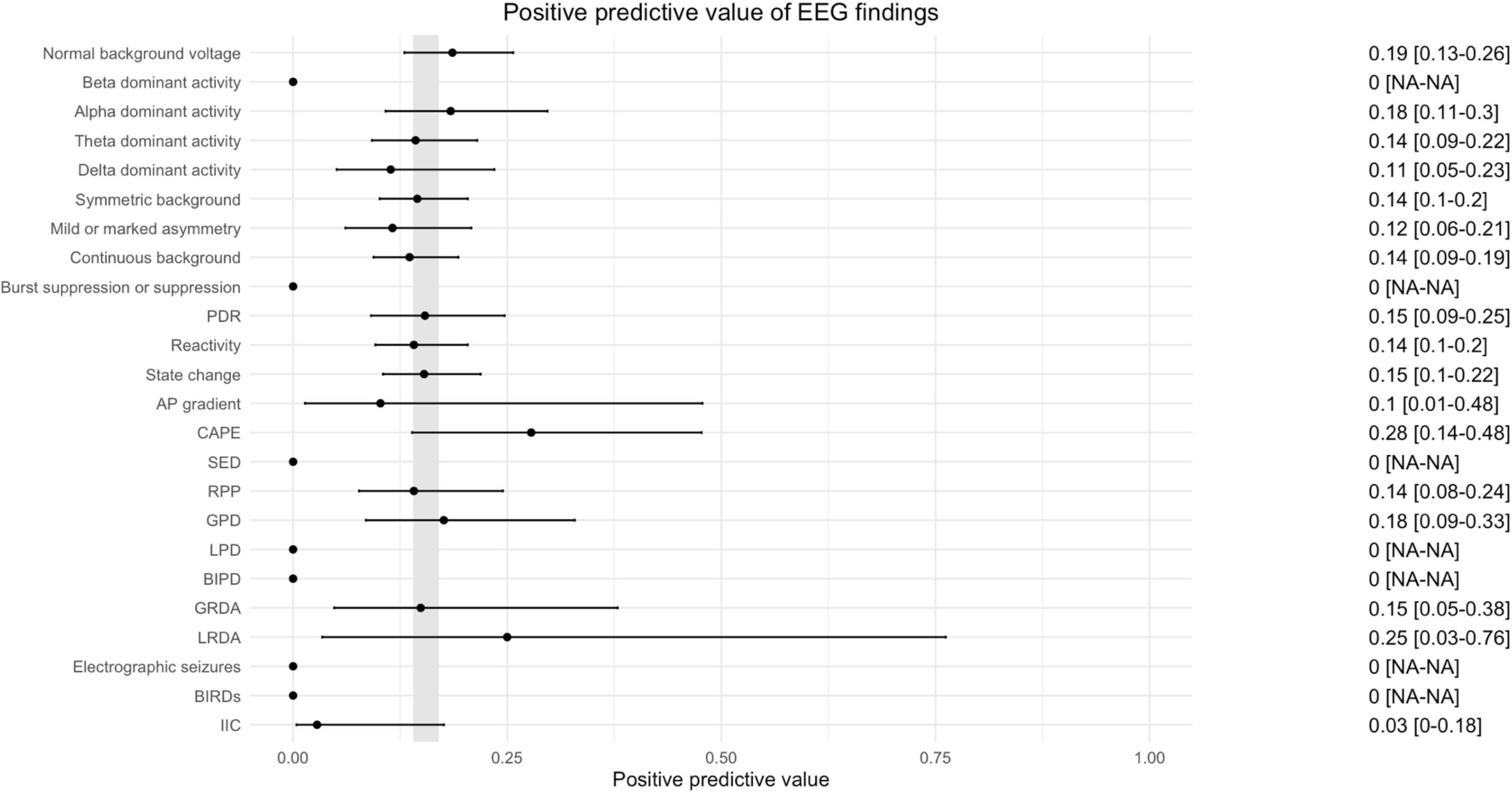
Positive predictive values of surface EEG findings for cognitive-motor dissociation. Bars represent 95% confidence intervals. Confidence intervals were not calculable (NA) for the positive predictive values of beta dominant activity, burst suppression or suppression, SED, LPD, BIPD, electrographic seizures and BIRDs due to no observed true positives. The gray shaded area in panel A represents the prevalence of CMD in the target population (14-17%).

**Figure 4.**
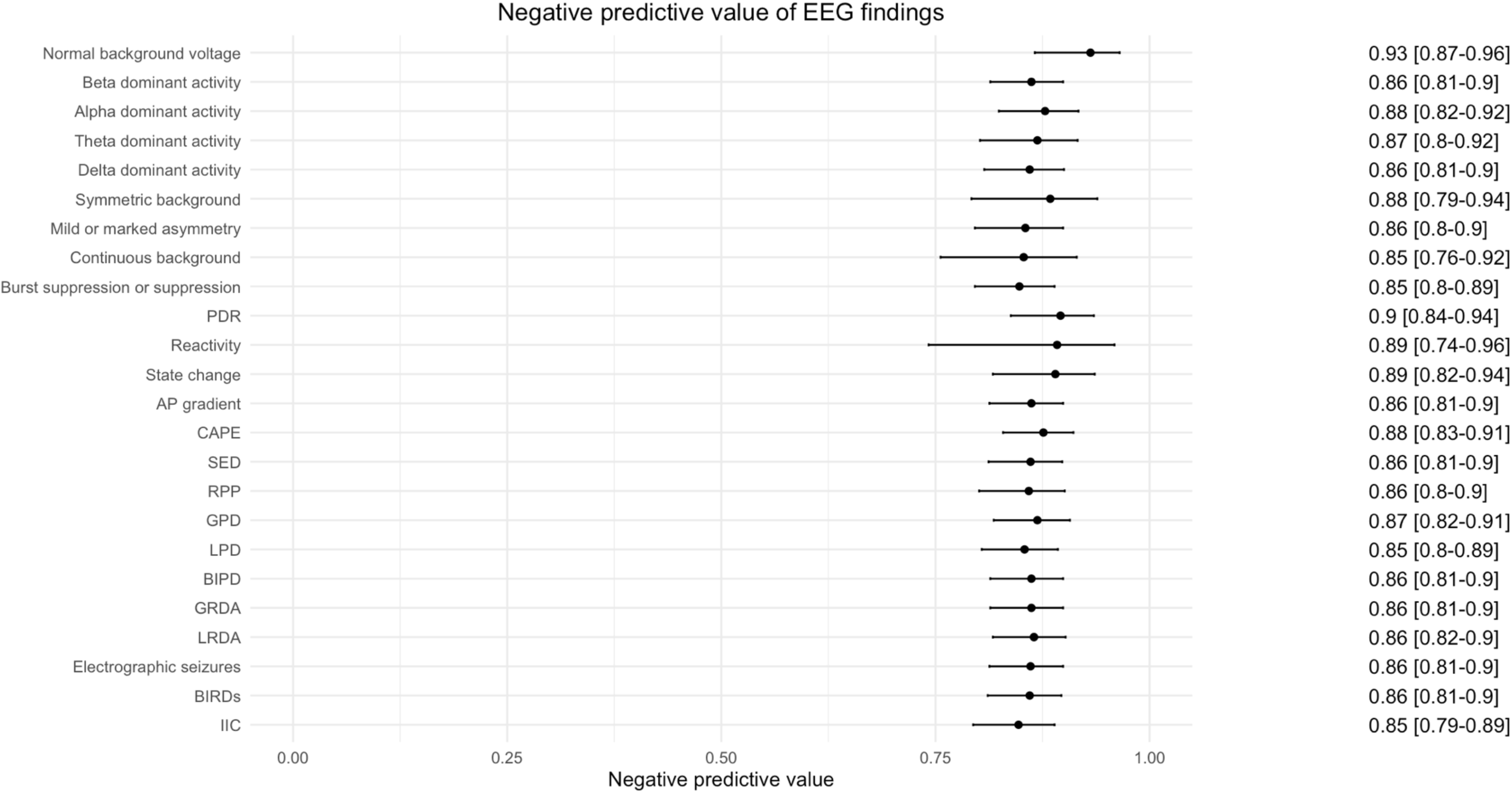
Negative predictive values of surface EEG findings for cognitive-motor dissociation. Bars represent 95% confidence intervals.

Normal background voltage, symmetry and continuity commonly co-occurred in surface EEGs, particularly in EEGs with a positive CMD test (**Figure 5**).

**Figure 5.**
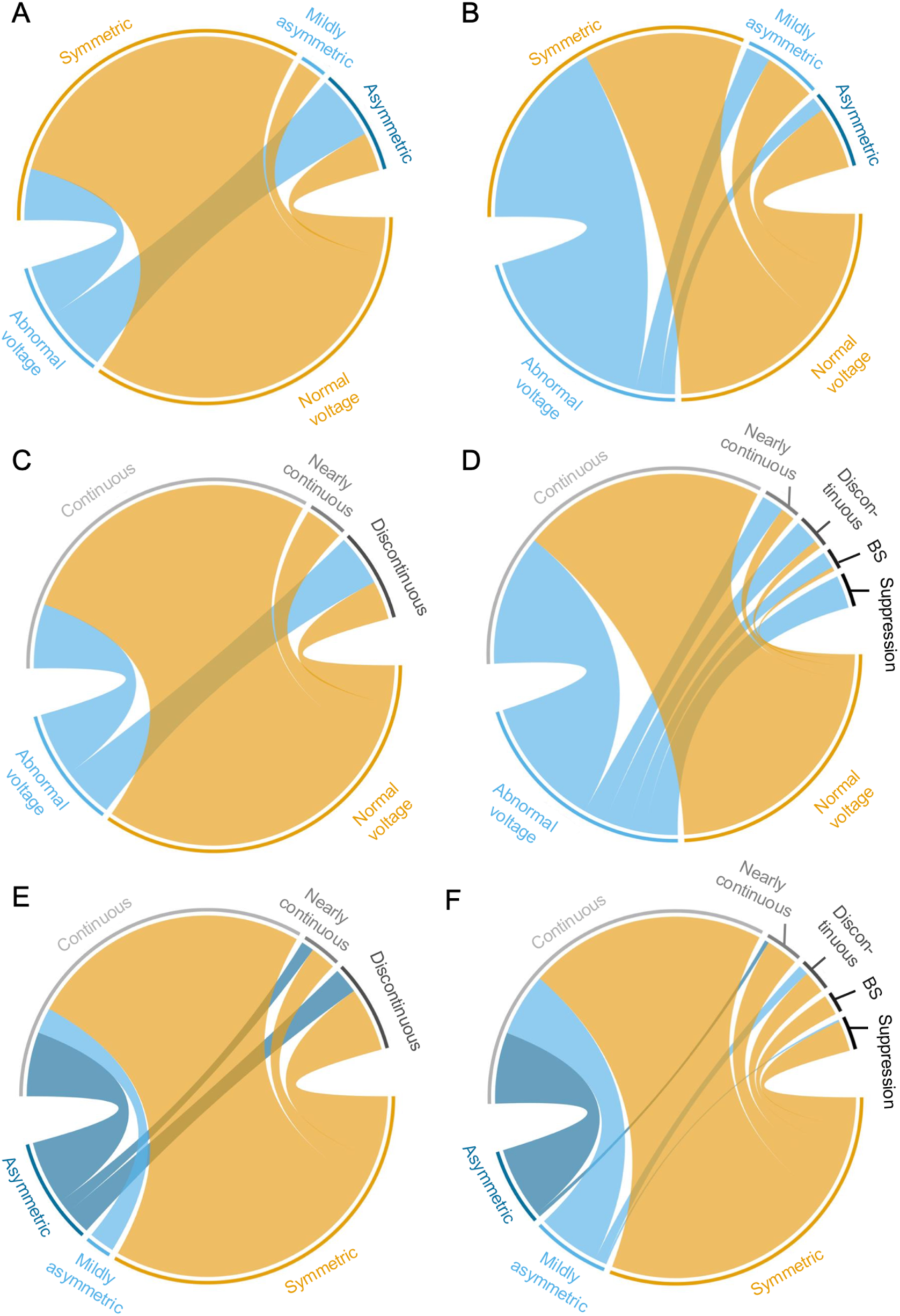
Chord diagrams illustrating the co-occurrence of EEG background activity characteristics. Chord diagram of voltage versus symmetry in EEGs with a positive (panel A) and a negative (panel B) CMD test. Chord diagram of voltage versus continuity in EEGs with a positive (panel C) and a negative (panel D) CMD test. Chord diagram of symmetry versus continuity in EEGs with a positive (panel E) and a negative (panel F) CMD test. BS denotes burst suppression.

### Inter-rater reliability

Inter-rater reliability results are provided in **Supplementary Table 3.** Concordance coefficients were assessed in 36 different EEG finding categories: they were perfect in 1 (3%), almost perfect in 10 (28%), substantial in 15 (42%), moderate in 6 (17%), fair in 3 (8%), slight in 1 (3%), and poor in 0 (0%).

## DISCUSSION

In this large prospective cohort study of critically ill patients unresponsive to verbal commands following an acute brain injury, we found that surface EEG findings are not reliable to screen for CMD or to identify patterns conferring higher CMD pretest probability. Normal EEG background voltage, symmetry and continuity are absent in one out of four positive CMD tests, such that the screening potential of these features is limited. Moreover, the PPV of these findings is close to CMD prevalence in the acute DoC population^5, 7, 8^: their presence on an EEG is therefore not informative for CMD pretest probability. We also found that all EEGs with burst suppression or suppression, SEDs, LPDs, BIPDs, electrographic seizures or BIRDs had negative CMD tests. Nevertheless, some of these findings were uncommon in our sample, such that their overall impact in avoiding negative CMD assessments may be modest.

A test’s PPV depends on its accuracy, namely its sensitivity and specificity, and the prevalence of the condition in the target population. In the present study, 14% of CMD tests were positive. In recent cohort studies on CMD prevalence and implications for long term prognosis, the proportion of clinically unresponsive patients with an acute brain injury who tested positive on CMD testing was 14-17%, which is in line with our findings.^5, 7, 8^ Our study does not address the accuracy of surface EEG findings for CMD in other pertinent patient populations, such as in individuals with chronic DoC in whom the prevalence of CMD is reported to be higher^5^.

In the critical care context, EEG monitoring is primarily used for detection of seizures but may also allow monitoring of cerebral blood flow, assessments of consciousness, and predictions of long term recovery.^17^ The EEG background can provide invaluable information into brain function of behaviorally unresponsive patients. Yet, the relationship between surface EEG findings and CMD status has received little attention to date.^9^ Although our study findings suggest that surface EEG has a limited role in optimizing CMD test allocation, future work could attempt to determine if certain combinations of EEG characteristics, in addition to other routinely available information, can more precisely discriminate patients at high CMD pretest probability. Studying the associations between normal EEG background activity characteristics and CMD, while adjusting for severity of acute brain injury and other potential confounders, could potentially shed light into the electrophysiological signature of CMD on surface EEG. Finally, whether certain EEG patterns interact with CMD in predicting recovery of consciousness and functional independence is unresolved and merits further investigation.

Our study has several strengths. We investigated the accuracy of surface EEGs in a large cohort of critically ill, brain injured, clinically unresponsive patients, which we enrolled prospectively and consecutively using a uniform set of selection criteria. We applied standardized ACNS critical care EEG criteria to optimize the validity and reliability of the index test. Electroencephalographers scoring EEGs were blinded to results from CMD testing, mitigating the risk of differential misclassification. All patients were exposed to the same verbal command paradigm during EEG acquisition, homogenizing their type and level of stimulation. Our study also has limitations. First, this was a single-center study involving adult patients, which limits the generalizability of our findings to other healthcare settings and the pediatric DoC population. Second, visual inspection of EEG is an inherently subjective process. Previous studies have shown difficulty in achieving good inter- and intra-rater reliability for some EEG features, such as reactivity.^18, 19^ However, by applying ACNS criteria, we used a widely accepted gold standard to classify ICU EEGs, and we measured at least substantial inter-rater reliability in >70% EEG features. We observed worse agreement for presence of a posterior dominant rhythm, anterior posterior gradient, and the ictal-interictal continuum, such that study results related to these patterns should be interpreted with caution. Blinding of electroencephalographers to CMD test results ensured misclassifications did not occur conditional to results from the reference standard, which could have introduced bias. Third, patients in the acute brain injury population frequently receive significant amounts of sedation, which can affect both surface EEG and CMD testing reliability. Should sedation cause differential misclassification in our study, our estimates may be biased. For instance, it is unknown whether the accuracy of medically induced burst suppression is similar to that of spontaneous burst suppression as seen in severe anoxic brain injury. Fourth, the exact accuracy and test-retest reliability of the reference standard in this study are unknown. There is currently no gold standard measurement for CMD, or covert consciousness overall, in the acute DoC population. Nevertheless, our EEG machine learning-based CMD detection algorithm based on a motor command paradigm has been shown to be clinically relevant due to its robust association with recovery of consciousness and recovery.^7, 8^ Finally, we excluded patients under high doses of sedation and patients with uncontrolled seizures since these individuals are assumed to have low CMD pretest probability. The EEG findings typically seen in these patients, such as epileptiform activity, seizures, and burst suppression or suppression backgrounds, are likely underrepresented in our sample due to these patient selection criteria.

In conclusion, surface EEG findings are not reliable to screen for CMD or to identify patterns conferring higher CMD pretest probability.

## ACKNOWLEDGEMENTS

We appreciate the assistance of the neuroscience ICU residents, fellows, and nurses.

## AUTHOR CONTRIBUTIONS

SE and JC designed the study. SE and EC performed surface EEG scoring. SE, JNB and VK performed data analysis. SE, JNB and JC drafted the manuscript. All other authors revised the manuscript for intellectual content. All authors reviewed and approved the final version of the manuscript. JC is the senior author and the guarantor of the study.

## DATA AVAILABILITY

The data that support the findings of this study are available from the corresponding author upon reasonable request.

