## Supplementary Material for "Surface EEG to identify cognitive motor dissociation after acute brain injury"

### TABLE OF CONTENT


Supplementary Figure 1 p. 2

Supplementary Table 1 p. 3

Supplementary Table 2 p. 4

Supplementary Table 3 p. 5

**Supplementary Figure 1.** Flowchart diagram.

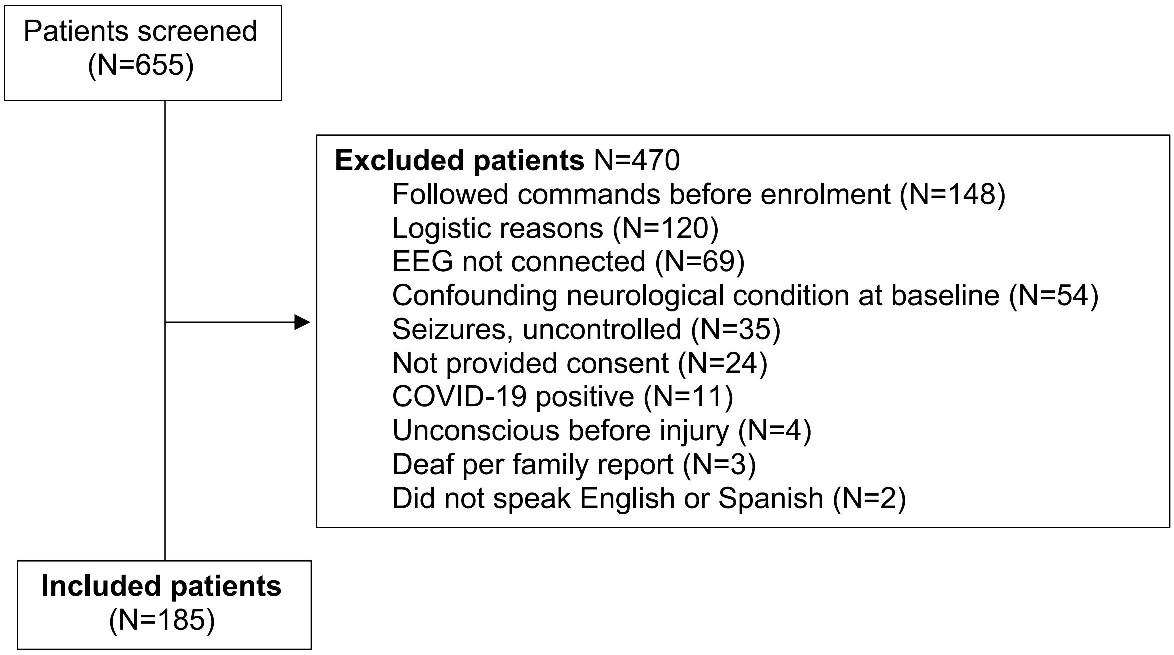


EEG denotes electroencephalogram.

**Supplementary Table 1.** Baseline characteristics of included and excluded patients.

| | Included (N=185) | Excluded (N=470) | OR (95% CI) | p-value |
|---|---|---|---|---|
| **Demographics** | | | | |
| Age < 63 years | 90 (49) | 234 (50) | 0.9 (0.7-1.3) | 0.74 |
| Female | 85 (46) | 234 (50) | 0.9 (0.6-1.2) | 0.36 |
| **Etiology** | | | | <0.001 |
| Intracerebral hemorrhage | 65 (35) | 89 (19) | Ref | |
| Cardiac arrest | 34 (18) | 80 (17) | 0.6 (0-26.6) | |
| Subarachnoid hemorrhage | 27 (15) | 33 (7) | 1.1 (0.5-2.7) | |
| Traumatic brain injury | 25 (14) | 42 (9) | 0.8 (0.2-2.8) | |
| Acute ischemic stroke | 14 (8) | 9 (2) | 2.1 (0.1-56.6) | |
| Other | 22 (12) | 215 (46) | 0.1 (0-215170.8) | |
| **Admission GCS < 8** | 148 (80) | 416 (89) | 0.3 (0.2-0.5) | <0.001 |

GCS denotes Glasgow Coma Scale.

**Supplementary Table 2.** Baseline characteristics of patients contributing one versus two EEGs.

| | One EEG (N=88) | Two EEGs (N=97) | OR (95% CI) | p-value |
|---|---|---|---|---|
| **Demographics** | | | | |
| Age < 63 years | 40 (45.5) | 50 (51.5) | 1.3 (0.7-2.3) | 0.40 |
| Female | 39 (44.3) | 46 (47.4) | 1.1 (0.6-2.0) | 0.67 |
| **Etiology** | | | | **0.01** |
| Intracerebral hemorrhage | 20 (22.7) | 45 (46.4) | Ref | 1 |
| Cardiac arrest | 18 (20.5) | 16 (16.5) | 0.4 (0.2-1.0) | |
| Subarachnoid hemorrhage | 12 (13.6) | 15 (15.5) | 0.6 (0.2-1.4) | |
| Traumatic brain injury | 12 (13.6) | 12 (12.4) | 0.5 (0.2-1.2) | |
| Acute ischemic stroke | 10 (11.4) | 3 (3.1) | 0.2 (0-0.6) | |
| Other | 16 (18.2) | 6 (6.2) | 0.2 (0.1-0.5) | |
| **Admission GCS < 8** | 62 (70.5) | 73 (75.3) | 0.8 (0.4-1.5) | 0.46 |

GCS denotes Glasgow Coma Scale.

**Supplementary Table 3**. Inter-rate reliability measures for surface EEG finding categories in the subsample of n=20 EEGs.

| Category [a] | Agreement | Cohen's kappa (95% CI) [b] |
|---|---|---|
| Symmetry | Almost perfect | 0.90 [0.73-1.00] |
| Frequency, right | Substantial | 0.75 [0.49-1.00] |
| Frequency, left | Almost perfect | 0.86 [0.68-1.00] |
| Posterior dominant rhythm, right | Moderate | 0.45 [0.22-0.67] |
| Posterior dominant rhythm, left | Fair | 0.31 [0.06-0.56] |
| Continuity, right | Substantial | 0.79 [0.57-1.00] |
| Continuity, left | Almost perfect | 0.81 [0.60-1.00] |
| Reactivity, right | Moderate | 0.42 [0.15-0.69] |
| Reactivity, left | Moderate | 0.48 [0.23-0.74] |
| State changes, right | Almost perfect | 0.82 [0.60-1.00] |
| State changes, left | Substantial | 0.72 [0.47-0.97] |
| Cyclic alternating pattern of encephalopathy, right | Perfect | 1.00 [1.00-1.00] |
| Cyclic alternating pattern of encephalopathy, left | Substantial | 0.64 [0.01-1.00] |
| Voltage, right | Substantial | 0.69 [0.41-0.97] |
| Voltage, left | Substantial | 0.62 [0.32-0.91] |
| Anterior posterior gradient, right | Slight | 0.20 [-0.25-0.65] |
| Anterior posterior gradient, left | Moderate | 0.51 [0.09-0.92] |
| Sporadic epileptiform discharges | Moderate | 0.52 [0.21-0.83] |
| Rhythmic periodic pattern, main term 1 | Almost perfect | 0.82 [0.58-1.00] |
| Rhythmic periodic pattern, main term 2 | Almost perfect | 0.91 [0.73-1.00] |
| Rhythmic periodic pattern, prevalence | Almost perfect | 0.91 [0.75-1.00] |
| Rhythmic periodic pattern, duration | Substantial | 0.74 [0.50-0.98] |
| Rhythmic periodic pattern, dominant frequency | Almost perfect | 0.82 [0.60-1.00] |
| Rhythmic periodic pattern, highest frequency | Substantial | 0.66 [0.42-0.91] |
| Rhythmic periodic pattern, phases | Substantial | 0.67 [0.31-1.00] |
| Rhythmic periodic pattern, sharpness | Substantial | 0.68 [0.32-1.00] |
| Rhythmic periodic pattern, absolute amplitude | Substantial | 0.80 [0.54-1.00] |
| Rhythmic periodic pattern, relative amplitude | Substantial | 0.71 [0.33-1.00] |
| Rhythmic periodic pattern, stimulus induced/terminated | Almost perfect | 0.81 [0.58-1.00] |
| Rhythmic periodic pattern, evolution pattern | Almost perfect | 0.90 [0.71-1.00] |
| Rhythmic periodic pattern, onset pattern | Fair | 0.27 [-0.01-0.54] |
| Rhythmic periodic pattern, triphasic | Moderate | 0.51 [0.03-0.98] |
| Rhythmic periodic pattern, lag | Substantial | 0.70 [0.31-1.00] |
| Rhythmic periodic pattern, polarity | Substantial | 0.62 [0.15-1.00] |
| Rhythmic periodic pattern, plus modifiers | Substantial | 0.80 [0.54-1.00] |
| Ictal-interictal continuum | Fair | 0.36 [-0.04-0.76] |

[a] None of the 20 EEGs had electrographic seizures, electroclinical seizures, electrographic status epilepticus, electroclinical status epilepticus, or brief potentially ictal rhythmic discharges (BIRDs).
[b] A value less than 0 signifies poor concordance; 0 to 0.20 indicates slight concordance; 0.21 to 0.40 suggests fair agreement; 0.41 to 0.60 denotes moderate

agreement; 0.61 to 0.80 reflects substantial agreement; and a range of 0.81 to 1.0 corresponds to almost perfect agreement.
